# Assessing the impact of optimized prevention strategies for mother-to-child HIV transmission dynamics in Kenya: a mathematical modeling study

**DOI:** 10.1101/2025.04.17.25326000

**Authors:** Robert Mureithi Maina, Samuel Musili Mwalili, Duncan Kioi Gathungu

## Abstract

HIV can be transmitted from a HIV infected mother to her child during pregnancy, delivery, or breastfeeding. According to NSDCC 2023, Kenya has estimated PMTCT coverage of 89.56% and PMTCT transmission rate of 8.6%. Even though there has been strides to address PMTCT, there is need to gear up approaches in addressing MTCT in order to significantly advance elimination. This research formulates a mathematical model to represent the dynamics of MTCT. Equilibrium points of the model are computed and the stability of HIV-free point is investigated. The numerical results show that a 50% decrease in maternal HIV transmission lowers infant infection rates by about 17.7%, whereas the same reduction in infant transmission decreases infections by nearly 39%, highlighting the greater sensitivity of infant transmission rates to direct interventions. While combination of strategies achieves the highest HIV minimization rates of up to 99.89% on infants, ART adherence alone significantly reduces transmission, particularly on infants (91.42%) while use of post-exposure prophylaxis (PEP) shows limited effectiveness when used alone(39.65%), suggesting that it should be complemented with other strategies for optimal impact. These findings emphasize the critical need for integrated interventions, where combining multiple prevention methods yields the best outcomes in reducing HIV infections on infants and moving closer to the elimination of pediatric HIV. These findings align with global recommendations from World Health Organization (WHO). This research can be used by the ministry of health to inform policy as well as recreated for other maternal infections.

**Author summary:** HIV can be transmitted from a mother to her child during pregnancy, delivery, or breastfeeding. In Kenya, despite efforts to prevent mother-to-child transmission (PMTCT), HIV transmission rates remain a concern. In this study, we developed a mathematical model to understand how HIV spreads from mothers to infants and to evaluate the effectiveness of different prevention strategies. Our findings highlight that reducing HIV transmission in mothers lowers infant infection rates, but direct interventions for infants, such as early antiretroviral therapy (ART) and post-exposure prophylaxis (PEP), have an even greater impact. A combination of strategies—ensuring mothers adhere to ART, providing PEP for infants, and promoting safe breastfeeding practices—was found to reduce HIV infections in infants by up to 99.89%. These results support the need for integrated approaches to HIV prevention. Policymakers and healthcare providers can use this research to refine HIV prevention programs, ensuring better maternal and infant health outcomes. Our model can also be adapted for other maternal infections, contributing to broader public health efforts in disease prevention.

## Introduction

HIV transmission from mother to their infants is a significant public health concern with profound biological, health, and social implications. HIV can be transmitted from an HIV-positive mother to her offspring through various biological mechanisms. During pregnancy, the virus can cross the placenta and infect the fetus, particularly in the later stages of gestation. During childbirth, the newborn can be exposed to HIV through contact with maternal blood and vaginal fluids. Additionally, HIV can be transmitted through breastfeeding, though the risk varies depending on factors like the viral load and breastfeeding practices of the mother (1).

Every year, roughly 1.3 million HIV-positive girls and women become pregnant, and if treatment is not received, the transmission rate to the unborn child ranges from 15% to 45%. (2). To address this, it is crucial to ensure immediate linkage to lifelong antiretroviral therapy (ART), ongoing care, and partner services upon HIV diagnosis. Although 85% of these women worldwide got access to ART by 2019, there are still issues with sustaining treatment and preventing transmission. The shift towards simplified, lifelong ART for pregnant women has made the eradication of HIV vertical transmission feasible. Global efforts, including the Triple Elimination Initiative, are focused on integrating these interventions into broader health services, enhancing disease monitoring, and promoting comprehensive sexual and reproductive health services. By 2030, WHO hopes to have eradicated the AIDS epidemic as a danger to public health (2).

PMTCT coverage in Kenya was 98% in Homa Bay County and 94% nationwide as of 2020 (3). Between 2015 and 2020, the national MTCT rate rose from 8.3% to 10.8%, which is a concerning trend. The risk of transmission is still very high during pregnancy and lactation. The National AIDS and STI Control Programme reports that 5% of pregnant women and 17% of nursing moms contract HIV during pregnancy and lactation, respectively, increasing the risk of HIV transmission to the fetus. Additionally, antiretroviral medication (ART) discontinuation rates among pregnant and lactating mothers with HIV are 47% and 21%, respectively, highlighting the increased risk of transmission and poor health outcomes during these periods. To achieve meaningful progress toward elimination, MTCT-addressing strategies must be given national priority and implemented.

The health effects HIV MTCT are serious and can manifest early in life. Infants infected with HIV may develop symptoms within the first few months, including failure to thrive, recurrent infections, and neurological issues. Without intervention, HIV-infected children can progress to AIDS, leading to severe opportunistic infections and early mortality. HIV infection also has long-term consequences, impairing the growth of a child, development, and overall health, impacting their cognitive and physical abilities (4)

Effective interventions exist to reduce the risk of HIV MTCT and improve outcomes for both mothers and infants (5). Antiretroviral therapy (ART) plays a vital part in prevention. Maternal ART, initiated during pregnancy and continued during breastfeeding if necessary, reduces the maternal viral load, significantly lowering the risk of transmission to the child. Administering ART to the newborn immediately after birth further reduces transmission risk and supports the health of child.

In cases where maternal viral load is not well controlled or other risk factors are present, a scheduled cesarean section may be recommended to reduce transmission during childbirth. Avoiding breastfeeding in settings where safe alternatives are available helps eliminate HIV transmission risk through breast milk (6). Infant prophylaxis with antiretroviral drugs is also recommended to reduce infection risk if breastfeeding is unavoidable or for other reasons (5).

Prevention strategies include routine HIV testing and counseling for pregnant women to identify infections early (7). Providing effective contraception to HIV-positive women who do not wish to conceive prevents unintended pregnancies and potential transmission. Complete care and assistance for mothers with HIV and their babies, including nutritional support and psychosocial counseling, are essential for optimizing health outcomes.

Some significant barriers hindering the adoption of elimination of MTCT services for women who have tested positive are issues with disclosure of HIV status (77.69%), dependency on partners (80.6%) and lack of assistance from family and partners (75%), long waiting times (33.3%), and lack of linkage to support groups (47.2%) (1). Effective implementation of tailored strategies can significantly enhance access, uptake, and retention in PMTCT programs, emphasizing the importance of local adaptation and addressing existing service gaps (8). There is need to an integration of healthcare services including ease for families by reducing HIV stigma and simplifying services (9).

From the previous research, various research gaps were identified. There is need to develop a mathematical model for mother-to-child HIV transmission and use the model to compose effective intervention combination strategies for MTCT using optimal control.

## Materials and methods

This section outlines the approach and methods used to achieve the objectives of the study. It begins by establishing the structure of the model, defining the key assumptions that guide its formulation, and representing the system through equations that describe its dynamics. The model is also analyzed for properties and stability. An optimal control framework is also presented. This section provides the necessary theoretical and computational foundation for evaluating the model’s outcomes.

### Model description

To describe the dynamics of MTCT in Kenya, a seven-state disease compartmental model is used to illustrate how mothers and infants move from one state to another. Every pregnant woman above 15 years who goes for ANC and has tested positive for HIV is under treatment. Children born by the infected mother are put on treatment in the 0-6 hours window after birth. Women with high viral load (*>* 1000 units) deliver through CS. The number of preterm babies is negligible. Delivery of twins, triplets or more babies is negligible. For the first several months following birth, infants are exclusively fed breast milk.

A population *A* of mothers come from ANC. A proportion *π*_*N*_ of them test negative for HIV to join HIV negative mothers class *M*_*N*_ . Another proportion *π*_*P*_ of *A* tests positive for HIV respectively to form HIV positive mothers class *M*_*P*_ . The rest are recruited to HIV suppressed mothers class,*M*_*S*_. *M*_*N*_ can acquire HIV at a rate *β*_0_ and join *M*_*P*_ . The *M*_*P*_ class can adhere to Art at a rate *α*_*am*_ and have their HIV viral load suppressed to join *M*_*S*_. *M*_*S*_ can drop out of ART at a rate *α*_*dm*_.

A proportion *π*_*N*_ + *kπ*_*P*_ + (1 *− π*_*p*_ *− π*_*N*_) of *A* or rather (1 + (*k −* 1)*π*_*P*_) *A* of the infants are recruited to HIV negative infants class, *I*_*N*_, where *k* is the proportion of negative infants born from *M*_*P*_ . Due to breastfeeding, HIV is transmitted by *M*_*P*_ to *I*_*N*_ at a rate *β*_1_ causing them to join the HIV exposed infants population, *I*_*E*_. This class is also added to it by a proportion 1 *− k* of infants from *M*_*P*_ . Post exposure prophylaxis is given to *I*_*E*_ infants and they regain their HIV negative status to go back to *I*_*N*_ at a rate *ρ*. At a rate *δ, I*_*E*_ population acquires HIV to join *I*_*P*_ class. ART adherence by *I*_*P*_ at a rate *α*_*ai*_ can lead them to join the HIV suppressed infants population, *I*_*S*_. ART dropout by *I*_*S*_ at a rate *α*_*di*_ leads to joining the *I*_*P*_ infants population. Mothers and children die at a natural death rate *µ*_*M*_ and *µ*_*I*_ respectively. Unfortunately, infants die due to HIV exposure are rate *d*_*I*1_ and high viral load of HIV at a rate *d*_*I*2_. Mothers also die due to HIV infection are rate *d*_*M*_ .

**MTCT strath.png**

### Model equations

Given the flow diagram in **??**, the parameter description in table 1, the non-linear ordinary differential equation system that follows is provided by,

**Table 1.**
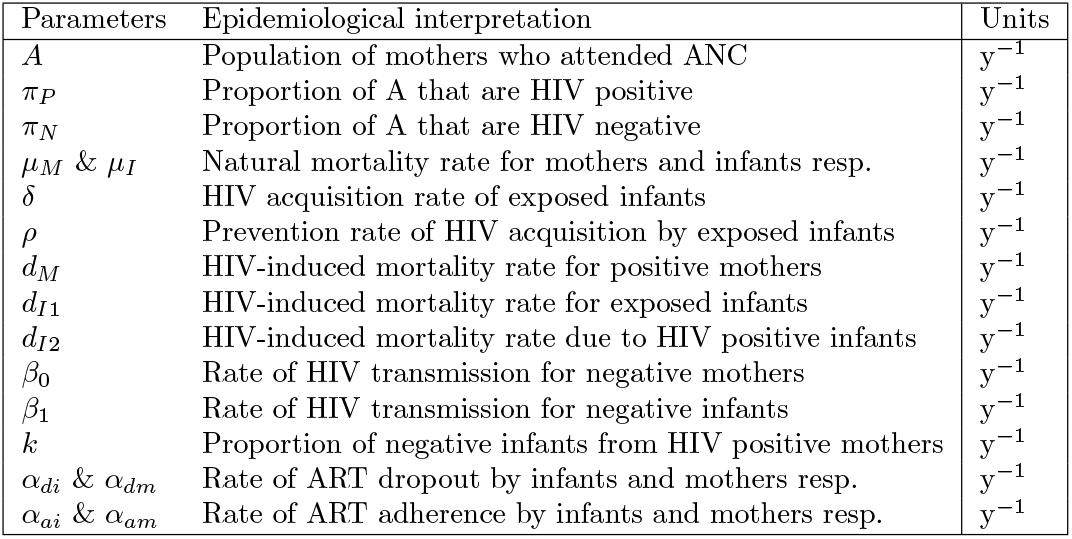
Parameters of the model; year(y).

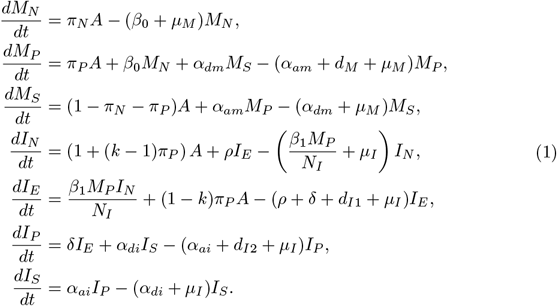

*N*_*I*_ = *I*_*N*_ + *I*_*E*_ + *I*_*P*_ + *I*_*S*_ represents the total population of infants.

### Properties of the model

Well-posedness of the model will be demonstrated. To prove the model’s well-posedness, the following have to be shown:

1. For the given initial conditions of the model (1), the solutions of our model system remains positive for all *t >* 0.
2. The solutions of a model’s system with the given initial conditions are bounded in a positive region,

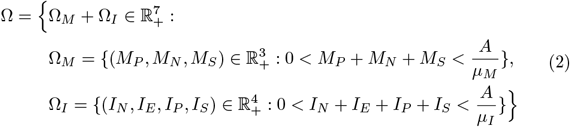

for any time *t ≥* 0.

**Positivity**

For realistic modeling of human population, all the state variables must be positive and the solutions to the model system with positive initial conditions should remain positive. This theorem is arrived at:

#### Theorem 1.

*For the given initial conditions of the model* (1), *the solutions of our model system remains positive for all t >* 0.

*Proof*. **HIV negative mothers**

Taking the equation for HIV Negative Women and considering the dynamics of *W*_*N*_ only,

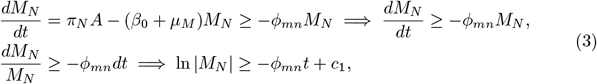

for constant *c*_1_ and *ϕ*_*mn*_ = (*β*_0_ + *µ*_*M*_). Taking the exponential for both sides of 3,

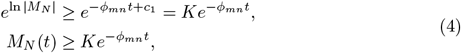

where 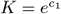 is a constant. Substituting the initial condition *M*_*N*_ (0) = *M*_*N*0_ in 4, then

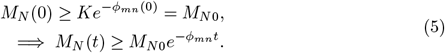

Hence *K* = *M*_*N*0_. The exponential part of 5 is always positive and *M*_*N*0_ *≥* 0, hence *M*_*N*_ (*t*) is always positive, meaning;

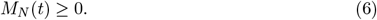

In a similar way, the procedure can be applied to all the remaining eight equations in model system (1), so that we have the following solutions,

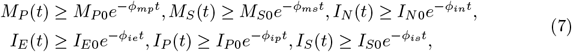

where 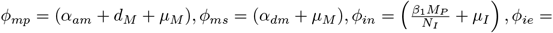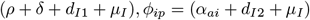 and *ϕ*_*is*_ = (*α*_*di*_ + *µ*_*I*_). Hence, all classes are **positive** given the initial conditions *M*_*P* 0_, *M*_*S*0_, *I*_*N*0_, *I*_*E*0_, *I*_*P* 0_, *I*_*S*0_.

### Boundedness

Boundedness ensures population sizes within each compartment cannot grow indefinitely or exceed reasonable and feasible range. This arrives at,

#### Theorem 2.

*The solutions of a model’s system with the given initial conditions are bounded in a positive region*,

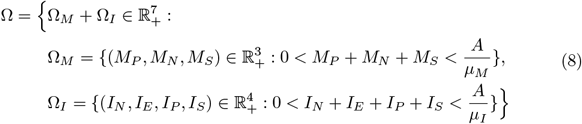

*for any time t ≥* 0.

*Proof*. Let *N*_*I*_ (*t*) = *I*_*N*_ (*t*) + *I*_*E*_(*t*) + *I*_*P*_ (*t*) + *I*_*S*_(*t*) and

*N*_*M*_ (*t*) = *W*_*N*_ (*t*) + *W*_*P*_ (*t*) + *M*_*P*_ (*t*) + *M*_*N*_ (*t*) + *M*_*S*_(*t*) be the total size of the population of infants and mothers. To show that the solutions of model system are bounded we proceed as follows:

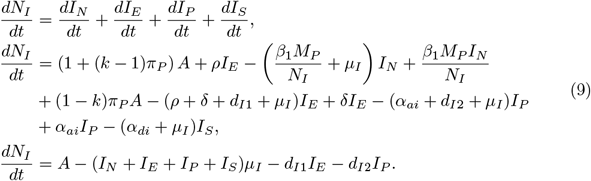

Since *N*_*I*_ = *I*_*N*_ + *I*_*E*_ + *I*_*P*_ + *I*_*S*_ and assuming there is no HIV induced mortality rate, (9) becomes,

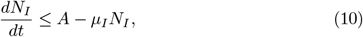

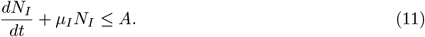

Using integrating factor *e*^*µ*^*I* to solve (11),

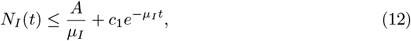

where *c*_1_ is a constant of integration. Applying the initial condition *N*_*M*_ (0) = *N*_*I*0_ in (12), we obtain,

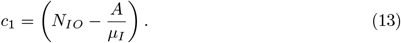

We then substitute the value *c*_1_ to *N*_*h*_(*t*) in (12) and simplify to get,

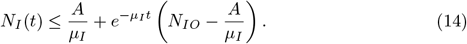

If 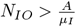, the right-hand side (RHS) of (14) experiences the largest possible value of *N*_*IO*_. That is, *N*_*I*_ (*t*) *≤ N*_*I*0_ for all *t >* 0. If 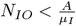, so that the largest possible value of the RHS of (14) approaches 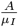 as time t goes to infinity. That is,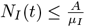 for all *t >* 0. Hence *N*_*I*_ (*t*) max 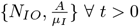, denoted as Ω_*I*_ .

The same is done for the mothers classes to get,

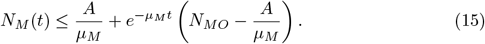

where *N*_*M*_ (*t*) = *M*_*P*_ (*t*) + *M*_*N*_ (*t*) + *M*_*S*_(*t*). If 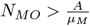, the right-hand side (RHS) of experiences the largest possible value of *N*_*MO*_. That is, *N*_*M*_ (*t*) *N*_*M*0_ for all *t >* 0.

If 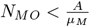, so that the largest possible value of the RHS of (15) approaches 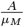 as time t goes to infinity. That is, 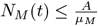 for all *t >* 0.

Hence 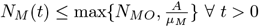, denoted as Ω_*M*_ .

### Stability analysis

#### HIV-Free equilibrium

The HIV-Free Equilibrium is obtained by setting the system of differential equations (1) to zero and setting all infected classes to zero. The *E*_*H*0_,

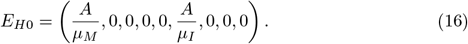

#### Endemic Equilibrium, *Ê* _*H*_

The EE equilibrium is obtained by setting the system of differential equations to zero and solving for each variable. The *E*_*H*_,

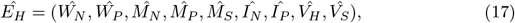

where,

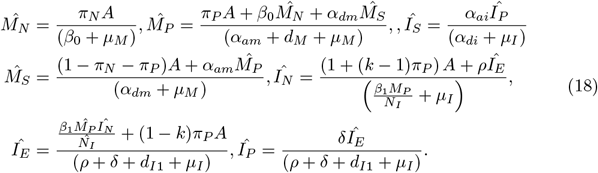

#### Local stability of HIV-Free equilibrium

The local stability analysis of the HIV-free equilibrium point (*E*_*H*0_) of the model is determined by finding the Jacobian matrix and its eigenvalues (10). An equilibrium point is locally asymptotically stable if all the eigenvalues of the Jacobian matrix at that point are negative. The general Jacobian matrix of model, 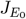 is given as:

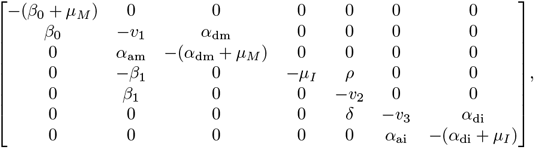

where *v*_1_ = *α*_am_ + *d*_*M*_ + *µ*_*M*_, *v*_2_ = *d*_I1_ + *δ* + *µ*_*I*_ + *ρ*, and *v*_3_ = *α*_ai_ + *d*_I2_ + *µ*_*I*_ . The eigenvalues of 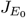 as solved by Mathematica software are,

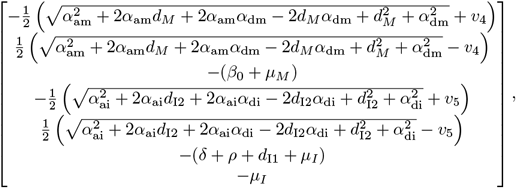

where *v*_4_ = *α*_am_ + *d*_*M*_ + *α*_dm_ + 2*µ*_*M*_ and *v*_5_ = *α*_ai_ + *d*_I2_ + *α*_di_ + 2*µ*_*I*_ . All the eigenvalues are negative except the second and seventh. Hence, *E*_0_ is locally asymptotically stable if,

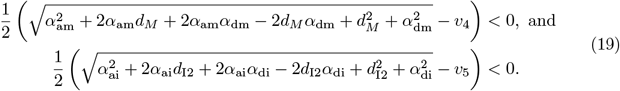

Simplifying (19) further and introducing arbitrary functions *ψ*_1_ and *ψ*_2_,

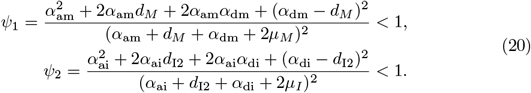

Hence, *E*_0_ is locally asymptotically stable if condition *ψ* = {*ψ*_1_, *ψ*_2_} in 20 is achieved.

#### Global stability of HIV-Free equilibrium

The method illustrated in (11; 12) is used to investigate the global asymptotic stability (GAS) of DFE point of the model, *E*_0_. Firstly, the model 1 must be written in the pseudotriangular form:

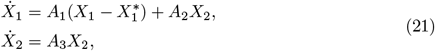

where *X*_1_ = (*M*_*N*_, *M*_*S*_, *I*_*N*_, *I*_*S*_), represents the number of uninfected and suppressed individuals and *X*_2_ = (*M*_*P*_, *I*_*E*_, *I*_*P*_) denotes the number of infected individuals. Let *X*^*\**^ be the HIV-free equilibrium. From *X*_1_,

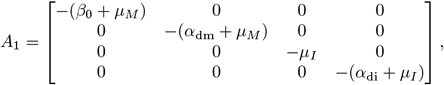

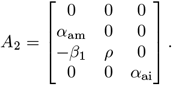

We can easily see that the eigenvalues shows that the subsystem *A*1are both real and negative. This shows that the subsystem 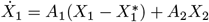, is globally asymptotically stable at the HIV free equilibrium 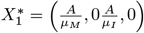. Additionally, from subsystem*X*_2_ = *A*_3_*X*_2_, we obtain the following matrix,

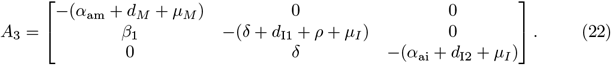

Notice that all the off-diagonal entries of *A*_3_ are nonnegative (equal to or greater than zero), showing that *A*_3_ is a Metzler matrix. To show the global stability of the HIV-free equilibrium *E*_0_, we need to show that the square matrix *A*_3_ in (22) is Metzler stable. We therefore need to prove the following;

##### Lemma 3.

*Let M be a square Metzler matrix that is block decomposed:*

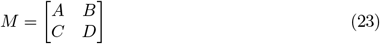

*where A and D are square matrices. The matrix M is Metzler stable if and only if A and D − CA*^*−*1^*B are Metzler stable*.

*Proof*. Matrix M in our case is *A*_3_. We therefore let,

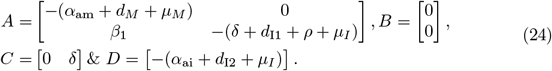

Clearly, A is Metzler stable.Then,

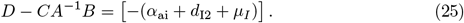

From (25), *D − CA*^*−*1^*B* is Metzler stable when the main diagonal element is strictly negative. Clearly, *D − CA*^*−*1^*B* is Metzler stable.

Thus the HIV free Equilibrium point *E*_0_ is globally asymptotically stable.

Epidemiologically, the above result implies that when there is no HIV infection, different human populations under consideration will stabilize at the *E*_0_. However, if there exists a HIV infection infection, then an appropriate control in forms of effective HIV treatment would be necessary to control the disease and restore the system to the stable HIV-free equilibrium.

### Sensitivity analysis formulation

The stability conditions derived at 20 can be used to establishing efficient control measures for the system 1. For its easiness to apply, Normalized Forward Sensitive Index method is used to determine the sensitivity indices as used in (13; 14). It’s index with regard to each parameter has been derived as follows to analyze the sensitivity of *ψ* = {*ψ*_1_, *ψ*_2_} to any parameter(say *µ*_*i*_),

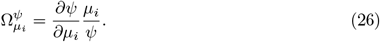

Where,

- 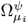 is positive, increase in *µ*_*i*_ leads to increase in *ψ* and,
- 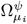 is negative, increase in *µ*_*i*_ leads to decrease in *ψ*.

The main goal of all control measures is to reduce the value of *ψ* = {*ψ*_1_, *ψ*_2_} and to analyze the propagation threshold such that effective interventions can be determined.

## Method of solution

**Runge Kutta method**, *𝒪* (*h*^4^)

The Runge-Kutta methods are designed to give greater accuracy and are more efficient in practical problems. They perform several function evaluations at each step and avoid the computation of higher order derivatives Runge-Kutta methods are known for their high accuracy. It is also stable and robust for stiff ODEs. It converges to the true solution, making it reliable for accurate approximations to the solution. These methods can be constructed for any order: second, third, fourth, fifth order, etc. The fourth order Runge-Kutta method is more popular.

Let consider the system described in equation 1 in its general form,

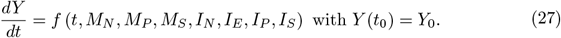

The fourth-order Runge-Kutta method for the system of equations in 27 is,

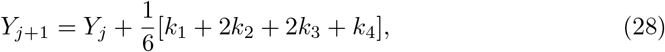

### Algorithm 1

Runge Kutta method,*O*(*h*^4^)

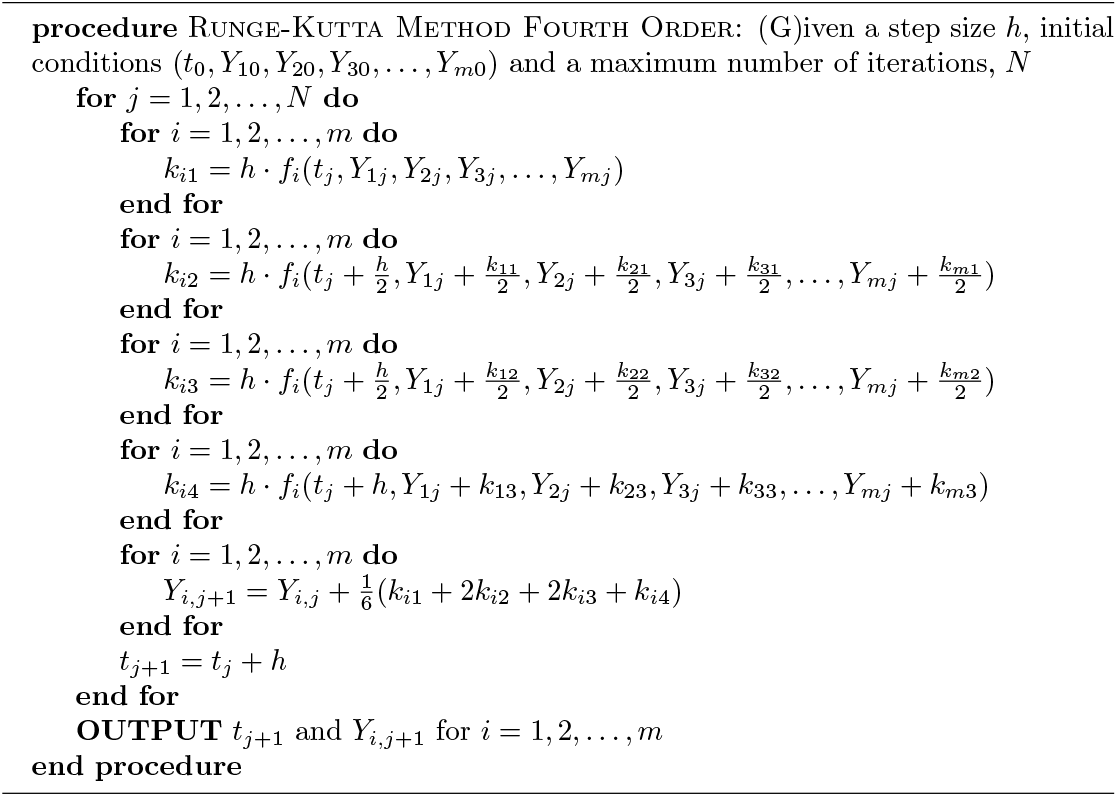

The error of this method is proportional to *h*^4^ and, therefore, can be improved by using small value of *h*.

### Optimal control model

In order to prevent mother-to-child transmission of HIV, the system (1) is extended into an optimal control problem by incorporating two time-dependent control functions.

These control functions are introduced at a specified time *t* with *t ∈* [0, *T*], as follows, where *T* is the final time.

1. *u*_1_(*t*) : **ART adherence**. For ART to suppress viral replication and remain effective over time, high levels of patient adherence are needed . It decreases vertical HIV transmission rates and significantly reducing maternal and infant morbidity and mortality (15).
2. *u*_2_(*t*) : **Post-exposure prophylaxis (PEP)**. The newborns receives a combination of Zidovudine (AZT) + Nevirapine (NVP) and/or Lamivudine (3TC) for 6–12 weeks (16). Nevirapine (NVP) prophylaxis protects infants from HIV infection during breastfeeding (17).

Including the control measures *u*_1_ and *u*_2_ in the model 1, the following optimal control model diagram is got,

#### MTCT strath2.png

The resulting equations from the optimal control model diagram **??** is,

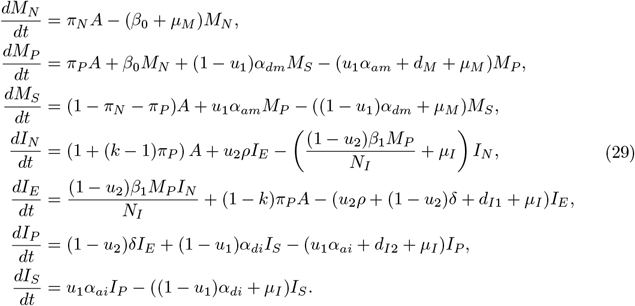

The initial conditions satisfy,

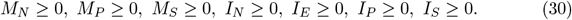

The Lebesgue measurable control set *U* is defined as follows in order to investigate the optimal control levels,

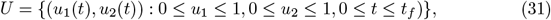

where *t*_*f*_ is the end time of implementing controls. The population of HIV positive mothers, HIV exposed infants and HIV positive infants is minimized by finding the optimal controls 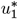 and 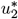 that leads to the following objective function,

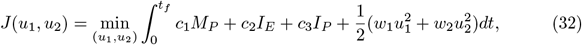

where *c*_1_, *c*_2_, *c*_3_, *w*_1_, and *w*_2_ are constants. Equations 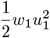 and 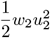 are the costs associated with the controls. The goal is to find the optimal controls 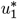 and 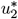 and optimal solutions by fixing the terminal time *t*_*f*_ that minimize the objective functional such that,

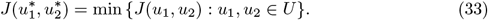

#### Existence of the optimal control

To show the existence of optimal control, the approach by (18) is used. It is already proved that the system (1) is bounded, so this result can be used to prove the existence of optimal control over finite time interval as applied in (18; HYPERLINK \l “bookmark40” 19). To ensure the existence of optimal control, following conditions must be checked if they are satisfied:

1. The set of controls and state variables be nonempty.
2. The control set *U* is convex and closed.
3. The right hand side of the state system is bounded by a linear function in the state and control variables.
4. The integrand of objective functional is convex on *U* .
5. The integrand of objective functional is bounded below by

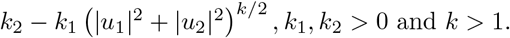

An existence of the state system with bounded coefficients has been used to give condition (*i*). The control set is convex and closed by definition hence (*ii*). The right hand side of the state system satisfies (*iii*). The state solutions are already bounded (*iv*). The integrand in the objective functional 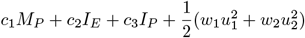 is clearly convex in *U* . For (*v*), from the bounds of the control system,

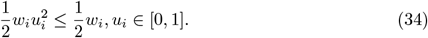

Also, considering the preceding inequality, the integrand can be written as

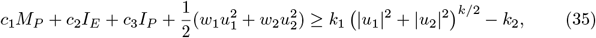

where 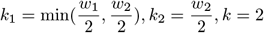. Therefore, there exists optimal control measures *u*_1_ and *u*_2_ that minimize the objective functional *J*(*u*_1_, *u*_2_).

#### The Hamiltonian and optimality system

The Pontryagins maximum principle stated the necessary conditions which are satisfied by optimal pair. Hence, by this principle, the Hamiltonian function (*H*) is obtained and defined as,

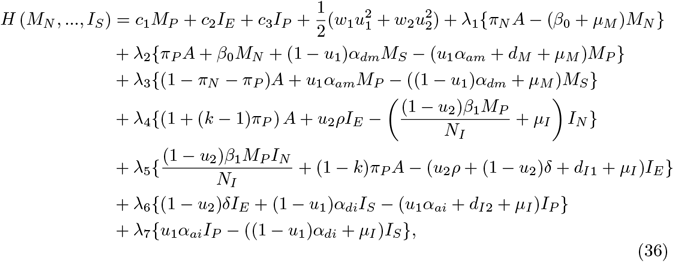

where,*λ*_*i*_, *i* = 1, …, 10 are the adjoint variables corresponding to state variables *M*_*N*_, *M*_*P*_, … and *I*_*S*_, respectively, and to be determined using Pontryagins maximal principle for the existence of optimal pairs.

##### Theorem 4.

*Let M*_*N*_, *M*_*P*_, *M*_*S*_, *I*_*N*_, *I*_*E*_, *I*_*P*_ *and I*_*S*_ *be optimal state solutions with associated optimal control variables u*_1_ *and u*_2_ *for the optimal control model, there exist co-state variables λ*_1_, …, *λ*_9_ *that satisfy*,

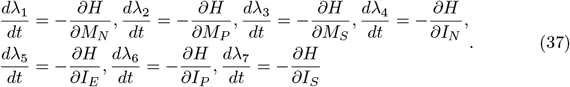

With transversality or final time conditions, *λ*_1_(*t*_*f*_) = … = *λ*_10_(*t*_*f*_) = 0, and where *H* is Hamiltonian function given in (*\**). Furthermore, the optimal controls 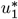, and 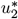 are,

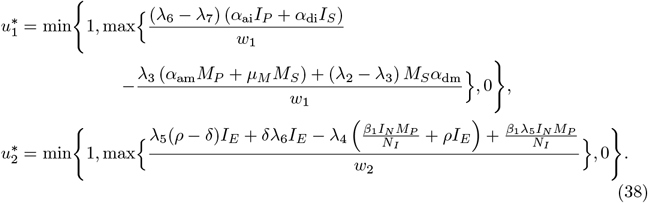

*Proof*. Pontryagins maximum principle gives the standard form of adjoint equation with transversality conditions (19). The standard results in (20) are applied to derive the adjoint relations, the transversality conditions and the optimal control system. Now, differentiating the Hamiltonian function with respect to state variables *M*_*N*_, *M*_*P*_, … and *I*_*S*_, respectively, the adjoint equations can be written as,

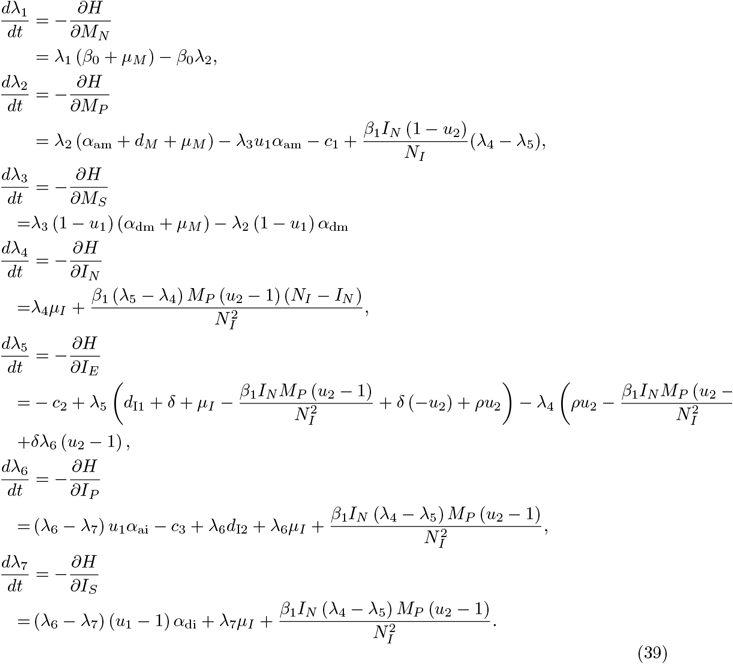

Further, the characterization of optimal controls 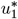, and 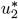 shows that,

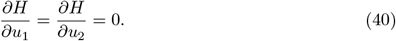

It follows that the optimal solution subject to constraints 0 *≤ u*_1_ *≤* 1, 0 *≤ u*_2_ *≤* 1 is,

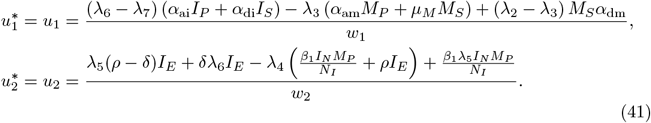

Using the equation (40), and the lower and upper bounds of four control measures, we obtained the characterization of optimal controls as follows.

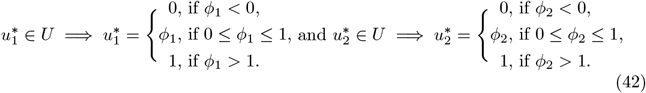

Where,

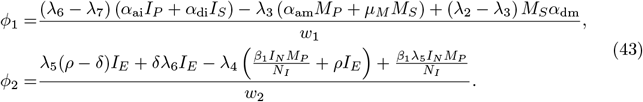

In compact form, the optimal controls can be written as,

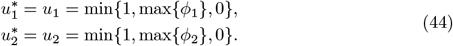

#### Optimal control algorithm

The Forward-Backward Sweep Method is a numerical technique often used to solve optimal control problems as seen in (21), it is an indirect method in which the differential equations from Pontryagin’s Maximum Principle are solved numerically. The idea of the Forward-Backward Sweep Method is that the state equations are solved forward in time, given an initial condition and the adjoint equations are solved backward in time given the value at final time. There are inherent difficulties in solving problems using indirect methods, however, the FBSM is easy to use on smaller problems and provides some insight into Pontryagin’s Maximum Principle.

##### Algorithm 2

Solving Optimal Control Problem using RK4 and Forward-Backward Sweep

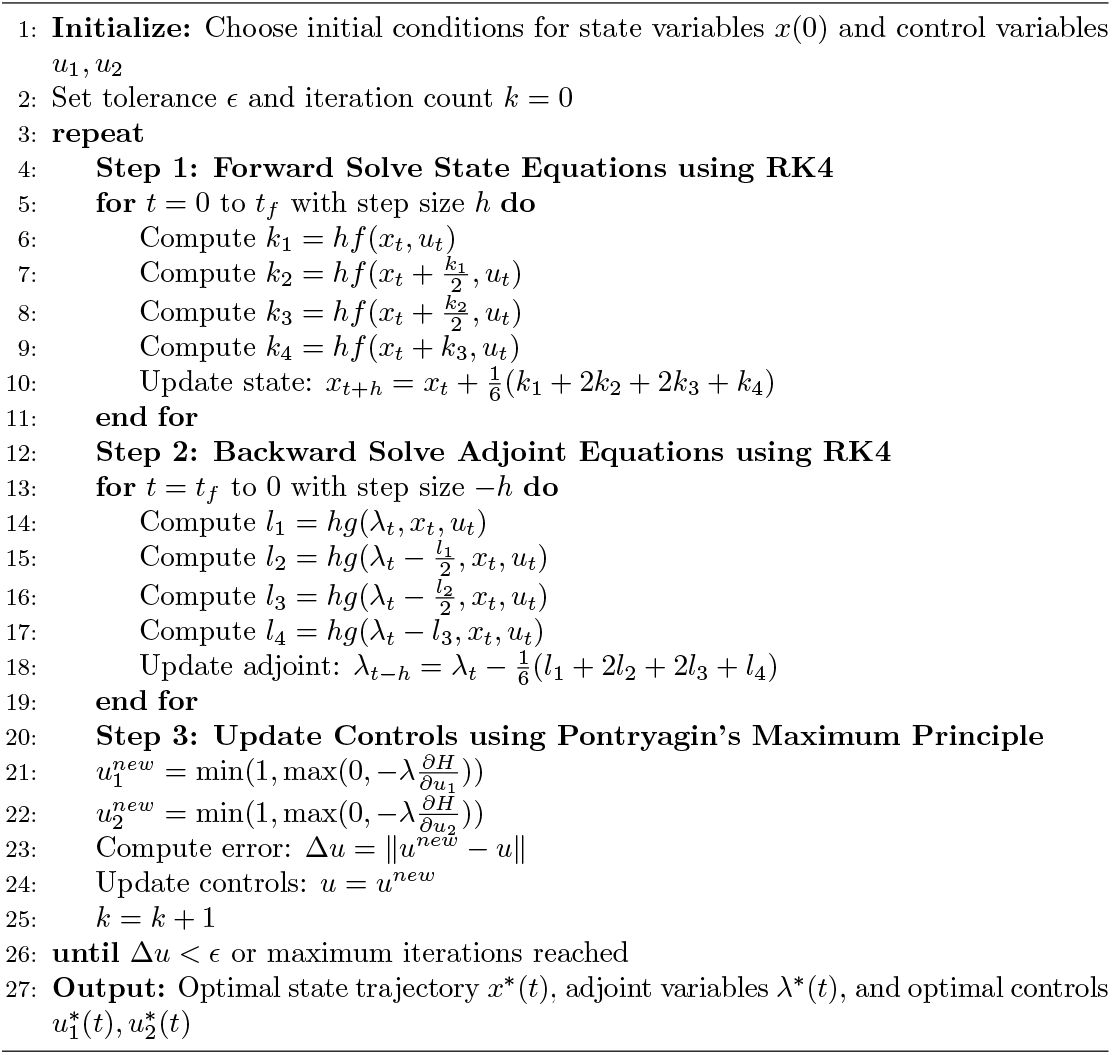

## Results

In this section, approximate solutions to the model equations 1 are computed using 𝒪 (*h*^4^) and 𝒪 (*h*^5^) order Runge-Kutta methods which are implemented via the solve ivp() function from Scipy library in Python. The initial populations are given by,

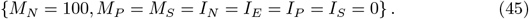

The simulation are run on time interval of [0, 100] years.

### Parameters

The parameter values are presented on table 2. They are arrived at from previous study sources and estimation.

**Table 2.**
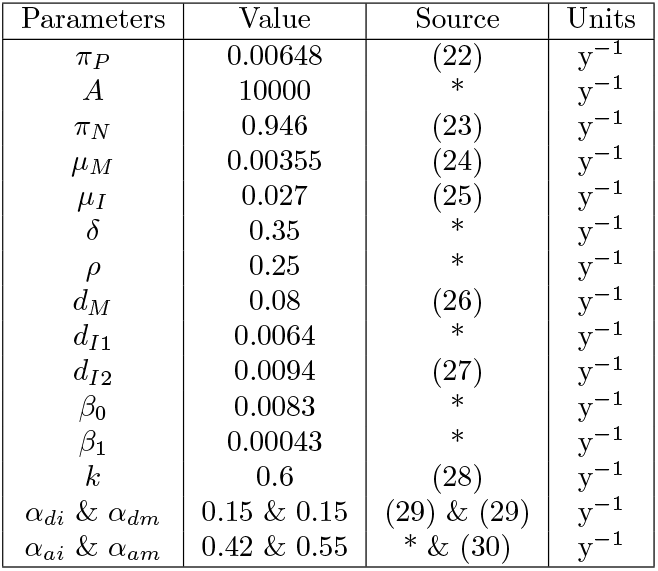
Parameters of the model; *year*^*−*1^(*y*^*−*1^).

| Parameters | Value | Source | Units |
| --- | --- | --- | --- |
| $\pi_P$ | 0.00648 | (22) | $y^{-1}$ |
| $A$ | 10000 | * | $y^{-1}$ |
| $\pi_N$ | 0.946 | (23) | $y^{-1}$ |
| $\mu_M$ | 0.00355 | (24) | $y^{-1}$ |
| $\mu_I$ | 0.027 | (25) | $y^{-1}$ |
| $\delta$ | 0.35 | * | $y^{-1}$ |
| $\rho$ | 0.25 | * | $y^{-1}$ |
| $d_M$ | 0.08 | (26) | $y^{-1}$ |
| $d_{I1}$ | 0.0064 | * | $y^{-1}$ |
| $d_{I2}$ | 0.0094 | (27) | $y^{-1}$ |
| $\beta_0$ | 0.0083 | * | $y^{-1}$ |
| $\beta_1$ | 0.00043 | * | $y^{-1}$ |
| $k$ | 0.6 | (28) | $y^{-1}$ |
| $\alpha_{di}$ & $\alpha_{dm}$ | 0.15 & 0.15 | (29) & (29) | $y^{-1}$ |
| $\alpha_{ai}$ & $\alpha_{am}$ | 0.42 & 0.55 | * & (30) | $y^{-1}$ |

where * =Assumed.

### Sensitivity analysis

In epidemic modeling, sensitivity analysis is performed to investigate model parameters with significant influence on a specific threshold and hence on the transmission and the spread of the disease under study (31). The model sensitivity analysis in this study is used to investigate parameter influence on the dynamics of HIV under different conditions on the stability thresholds *ψ* = {*ψ*_1_, *ψ*_2_}. In order to eliminate the HIV, the stability threshold number should be less than one, that is, *ψ <* 1.

From table 3, a positive sign on the SI indicates that an increase in the value of such a parameter increases the value of *ψ* = {*ψ*_1_, *ψ*_2_} and hence the growth of infection. On the other hand, a negative sign is indicative of a parameter that negatively affects *ψ* = {*ψ*_1_, *ψ*_2_}.

**Table 3.** Sensitivity indices.

| Parameters | Sensitivity index | Description |
| --- | --- | --- |
| $\mu_M$ | -0.0180409 | $\mu_M \sim \frac{1}{\psi}$ |
| $\mu_I$ | -0.170508 | $\mu_I \sim \frac{1}{\psi}$ |
| $d_M$ | -0.0662329 | $d_M \sim \frac{1}{\psi}$ |
| $d_{I2}$ | -0.0137669 | $d_{I2} \sim \frac{1}{\psi}$ |
| $\alpha_{di}$ | 0.0359026 | $\alpha_{di} \sim \psi$ |
| $\alpha_{dm}$ | -0.0492402 | $\alpha_{dm} \sim \frac{1}{\psi}$ |
| $\alpha_{ai}$ | 0.148373 | $\alpha_{ai} \sim \psi$ |
| $\alpha_{am}$ | 0.133514 | $\alpha_{am} \sim \psi$ |

### Graphs and discussions

In order to illustrate the feasibility of the theoretical results and the control strategies, graphs emanating from the numerical simulations are given. The python library, Matplotlib, is used to make the plots showing mothers and infants population dynamics over time.

#### Normal dynamics

The system 1 is solved numerically with method described in 1 and the graphs are presented as follows.

The graph on Fig **??** presents the trends of different maternal populations concerning HIV status over time. The HIV-negative mothers exhibit a rapid increase and stabilize at at 45 years, indicating effective preventive measures in reducing new infections. These trends align with studies such as (28), which highlight the success of prevention programs in reducing maternal HIV transmission. The HIV-suppressed group (green) also rises significantly before leveling off, demonstrating the impact of ART in controlling viral load among infected mothers. The HIV-positive mothers (red) population grows initially but stabilizes at a lower level due to ART interventions and mortality effects which emphasize the benefits of early ART in improving maternal health outcomes (32).

**mothers.png**

Fig **??** illustrates the dynamics of different infant populations in relation to HIV transmission, exposure and suppression. The HIV-negative infant population exhibits a sharp increase upto the 10^*th*^ year before stabilizing at equilibrium, indicating effective prevention measures. The HIV-suppressed infants population follows a rapid rise and levels off below the HIV-negative group, highlighting the impact of early ART initiation. The HIV-positive and HIV-exposed infants populations reach a steady state, reflecting the balance between new infections, treatment, and mortality. These trends align with findings from studies such as (33), which demonstrated the role of maternal interventions in reducing mother-to-child transmission, and (32), which emphasized early ART in improving infant survival rates.

**infants.png**

In order to control HIV transmission to mothers and infants, various prevention measures have to be undertaken. Based on model 1, we reduce the transmission rates by investigating the effects of varying the following parameters,

1. **HIV prevention among mothers**, *β*_0_: HIV transmission to mothers can be reduced by regular HIV testing, pre-exposure prophylaxis (PrEP) for high-risk mothers, safe sex practices (Use of condoms, limit sexual partners), voluntary medical male circumcision for the male spouses and ensuring safe blood transfusions and medical procedures (34). After varying the parameters identified above, the following results are achieved. From Fig **??** and **??**, the HIV transmission rate to mothers is lowered by about 30% *⇒ β*_0_ = 0.00581 and 50% *⇒ β*_0_ = 0.00415: This causes the infants infected with HIV to reduce by about 9% and 17% respectively. **mothers_posi_vary.png** **infants_vary1.png**
2. **HIV prevention among infants**, *β*_1_ & *k*: HIV prevention in infants involves maternal ART, infant antiretroviral prophylaxis, safe delivery and feeding practices, and early HIV testing to reduce the risk of transmission (9). After varying the parameters identified above, the following results are achieved. From Fig **??**, the HIV transmission rate to infants is lowered by about 30% *⇒ β*_1_ = 0.000301 & *k* = 0.78 and 50% *⇒ β*_0_ = 0.000215 & *k* = 0.9: This causes the infants infected with HIV to reduce by about 22% and 39% respectively. **infants_vary2.png**

The results on table 4 show that reducing HIV transmission in mothers has a notable impact on preventing infections in infants, but direct reductions in infant transmission have an even greater effect. A 50% decrease in maternal transmission (mothers) lowers infant infection rates by about 17.7%, whereas the same reduction in infant transmission decreases infections by nearly 39%, highlighting the greater sensitivity of infant transmission rates to direct interventions. While lowering maternal transmission remains crucial, targeted infant interventions, such as prophylaxis and early ART, yield significantly higher reductions in MTCT. These findings are of great health significance of optimizing prevention programs (9; 33), as even modest reductions in transmission rates can lead to major public health benefits, ultimately aiding in the elimination of pediatric HIV.

**Table 4.** Summary table. Effects of varying transmission parameters summary(%): MP - HIV positive mothers, IE - HIV exposed infants and IP - HIV positive infants.

| Varying | MP | IE | IP |
| --- | --- | --- | --- |
| 30% decrease in HIV transmission to mothers | −12.57% | −8.55% | −8.6% |
| 50% decrease in HIV transmission to mothers | −24.90% | −17.67% | −17.77% |
| 30% decrease in HIV transmission to infants | — | −21.50% | −21.52% |
| 50% decrease in HIV transmission to infants | — | −39% | −39.02% |

#### Optimal control dynamics

The system 29 is solved numerically to create simulation graphs presented as follows. The curve in the Fig **??** compares the population dynamics of HIV-positive mothers under two scenarios: with and without interventions. Without interventions, the number of HIV-positive mothers rapidly increases and stabilizes after infecting a high population. However, with ART adherence and PEP in place, the population of HIV-positive mothers remains significantly lower by 35.94%, demonstrating the effectiveness of these controls in reducing HIV prevalence. (35) highlights the crucial role of HIV medicine in minimizing maternal HIV burden and improving health outcomes.

#### mothers_posi_with.png

Fig **??** illustrates the population dynamics of HIV exposed and HIV positive infants population under HIV control strategies and without the strategies. The first set of graphs shows the natural progression of HIV-exposed infants population, where it later stabilizes. In contrast, with interventions, a significant reduction of 60.60% in the number of HIV-exposed infants is realized, indicating the effectiveness of ART adherence and PEP uptake in preventing mother-to-child HIV transmission. HIV positive infants population represented by the log-scale also reduces by 97.82% with control strategies. The logarithmic scale is used to magnify the results since the controls greatly minimize the exposed infants. This reinforces findings from (5) which show the impact of ART adherence and PEP in reducing vertical transmission rates and long-term HIV prevalence.

#### infants_all_with.png

Fig **??** compares the impact of ART adherence and post-exposure prophylaxis (PEP) on the population of HIV-positive mothers over time. Without any control measures, high population of mothers remain HIV-positive. ART adherence alone, leads to a significant reduction of 85.14% in the HIV-positive mothers’ population over time. Notably, PEP doesn’t have effect in reducing the HIV positive mothers population since in this model PEP is only focused on infants. This effects the critical role of ART adherence in reducing maternal HIV prevalence and preventing transmission to infants (7).

#### mothers_comp.png

Fig **??** compares the distinct effect of ART adherence and post-exposure prophylaxis (PEP) on the population of HIV-exposed and HIV-positive infants over time. Without control measures, the populations grows substantially. ART adherence shows a significant reduction in HIV exposure among infants by 65.78% and much more on HIV positive infants by 91.42%. PEP also leads to a reduction, though not as substantial as ART adherence with HIV exposure among infants reducing by 13.28% and HIV positive infants by 39.65%. These results show that ART adherence is highly effective in reducing mother-to-child HIV transmission, while PEP provides an additional layer of prevention when ART adherence is incomplete.

#### infants_all_comp.png

The graph in Fig **??** compares the population of HIV-positive mothers under two scenarios: without optimal control and with combined strategies of ART adherence and PEP. The results indicate that without intervention, the population of HIV-positive mothers grows significantly over time, stabilizing at a high steady-state . However, the application of both ART adherence and PEP controls substantially reduces the number of HIV-positive mothers by 66.24%, emphasizing the effectiveness of comprehensive HIV management strategies. These findings align with studies highlighting the impact of ART in reducing maternal HIV transmission and PEP in preventing new infections, reinforcing the necessity of integrated control measures to curb HIV spread among mothers (36).

#### mothers_posi_comb.png

Fig **??** illustrates the population dynamics of HIV-exposed infants and contrasts without optimal control and with combined strategies of ART adherence. The results reveal that in the absence of intervention, the number of HIV-exposed infants rises sharply before stabilizing at the equilibrium. However, the synergy of ART adherence and PEP significantly reduces the number of HIV-exposed infants by 93.28%, demonstrating the effectiveness of these interventions in preventing mother-to-child transmission (PMTCT). This shows that ART is critical in reducing vertical HIV transmission and PEP in mitigating post-exposure risks.

#### infants_exp_comb.png

Investigating HIV-positive infants population on Fig **??** on the two scenarios: without optimal control and with combined strategies of ART adherence and PEP, interesting results are found. In the absence of intervention, the number of HIV-positive infants grows exponentially before reaching the equilibrium, reflecting a continuous transmission from mother-to-child. However, when ART adherence and PEP are implemented together, the HIV-positive infant population remains near zero(reduces by 99.89%), highlighting the effectiveness of these strategies in eliminating vertical transmission. The log scale emphasizes that, even at early stages, intervention significantly curtails the exponential rise of new HIV-positive cases. These findings are consistent with (37) report, which emphasize that a combination of ART adherence and PEP can reduce the risk of perinatal HIV transmission to below 5%, significantly improving infant health outcomes.

#### infants_posi_all_comb.png

The results in Table 5 highlight the effectiveness of various optimal control strategies in reducing mother-to-child transmission (MTCT) of HIV, with the combination of strategies achieving the highest minimization rates of up to 99.89% in infants. ART adherence alone significantly reduces transmission, particularly in mothers (88.14%), reinforcing the importance of sustained antiretroviral therapy in prevention programs. The use of post-exposure prophylaxis (PEP) shows limited effectiveness when used alone, suggesting that it should be complemented with other strategies for optimal impact. These findings emphasize the critical need for integrated interventions, where combining multiple prevention methods yields the best outcomes in reducing HIV infections in infants and moving closer to the elimination of pediatric HIV.

**Table 5.**
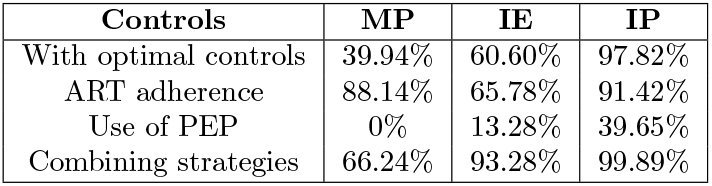
Summary table. Optimal control graphs summary(%): MP - HIV positive mothers, IE - HIV exposed infants and IP - HIV positive infants.

**Table 6.** Abbreviations.

| Abbreviation | Meaning |
| --- | --- |
| AIDS | Acquired Immunodeficiency Syndrome |
| ANC | Antenatal Care |
| ART | Anti Retroviral Therapy |
| CLHIV | Children Living with HIV |
| HIV | Human Immunodeficiency Virus |
| MTCT | Mother To Child Transmission |
| PEP | Post-Exposure Prophylaxis |
| PMTCT | Prevention of MTCT |
| STI | Sexually Transmitted Infections |
| VLS | Viral Load Suppression |
| WHO | World Health Organization |
| $M_N$ | HIV negative mothers |
| $M_P$ | HIV positive mothers |
| $M_S$ | Mothers with suppressed HIV viral load |
| $I_P$ | HIV positive infants |
| $I_E$ | HIV exposed infants |
| $I_N$ | HIV negative infants |
| $I_S$ | Infants with suppressed HIV viral load $VL < 50c/ml$ |

## Conclusion

In this research, a mathematical model representing the dynamics of mother-to-child HIV transmission and optimal control is developed. This study considered two populations; mothers and infants. This study proved that the formulated model is biologically and mathematically well posed on an invariant region Ω. HIV-free equilibrium is shown to be locally asymptotically stable by use of Jacobi method. The global stability of the HIV-free equilibrium is only guaranteed if the threshold quantity *ψ* = {*ψ*_1_, *ψ*_2_} is less than unity.

The numerical results emphasize the significant impact of reducing HIV transmission rates through targeted interventions, particularly in preventing mother-to-child transmission (MTCT). A reduction in transmission rates to mothers and infants leads to a significant decline in pediatric HIV cases, with the greatest benefits seen when multiple strategies are combined. A 30% to 50% reduction in maternal HIV transmission notably decreases infection rates in infants, reinforcing the necessity of early intervention and maternal treatment during pregnancy and breastfeeding. These findings highlight the importance of prevention programs, as even modest reductions in transmission rates can result in substantial public health benefits.

Furthermore, the effectiveness of optimal control strategies, particularly the combination of ART adherence, post-exposure prophylaxis (PEP), and other interventions, demonstrates the need for integrated healthcare approaches. The combination of strategies achieves the highest transmission reduction, nearing complete prevention (99.89%), proving that a multi-faceted approach is essential in eliminating pediatric HIV. While ART adherence alone shows a high impact, it is clear that a comprehensive prevention plan yields the best results. These conclusions support the need for sustained policy implementation, increased ART access, and community awareness to ensure effective HIV prevention and control in maternal and child health programs.

ART adherence is highly effective in reducing mother-to-child HIV transmission, while PEP provides an additional layer of prevention when ART adherence is incomplete. However, the combined strategy of ART adherence and PEP achieves the most significant reduction, demonstrating the importance of integrating multiple prevention approaches. These findings align with global recommendations from World Health Organization (WHO).

Future studies could incorporate more comprehensive models that account for additional factors such as socioeconomic status, adherence variability, and drug resistance in HIV prevention strategies. Empirical validation of the mathematical findings using clinical and epidemiological data through fitting could improve the accuracy and applicability of the study’s conclusions. Utilizing AI-driven predictive models could enhance the precision of intervention strategies and optimize control measures in diverse populations. Health policy makers e.g. the ministry of health should strengthen ART adherence programs, expand PEP access for infants, and implement integrated prevention strategies combining ART, PEP, education, and regular screening. This study recommends investment in maternal and child healthcare infrastructure, ensure routine monitoring and evaluation of prevention programs, and promote community education on early testing and HIV prevention.

Abbreviations

The abbreviations used in the article as well as the model population classes are presented below.

## Author contributions

**Conceptualization:** Robert Mureithi Maina, Samuel Musili Mwalili and Duncan Kioi Gathungu.

**Data curation:** Robert Mureithi Maina.

**Formal analysis:** Robert Mureithi Maina, Samuel Musili Mwalili and Duncan Kioi Gathungu.

**Investigation:** Robert Mureithi Maina, Samuel Musili Mwalili and Duncan Kioi Gathungu.

**Methodology:** Robert Mureithi Maina, Samuel Musili Mwalili and Duncan Kioi Gathungu.

**Visualization:** Robert Mureithi Maina.

**Writing original draft:** Robert Mureithi Maina.

**Supervision**: Samuel Musili Mwalili and Duncan Kioi Gathungu.

**Reviewing and editing:** Robert Mureithi Maina, Samuel Musili Mwalili and Duncan Kioi Gathungu.

## Data availability statement

All code used for simulation and plotting is available on a GitHub repository at Github.

## Acknowledgments

We are very grateful to the Centre for Health Analytics and Modelling (CHAM) Strathmore University for their support and collaboration throughout the duration of this study.

---

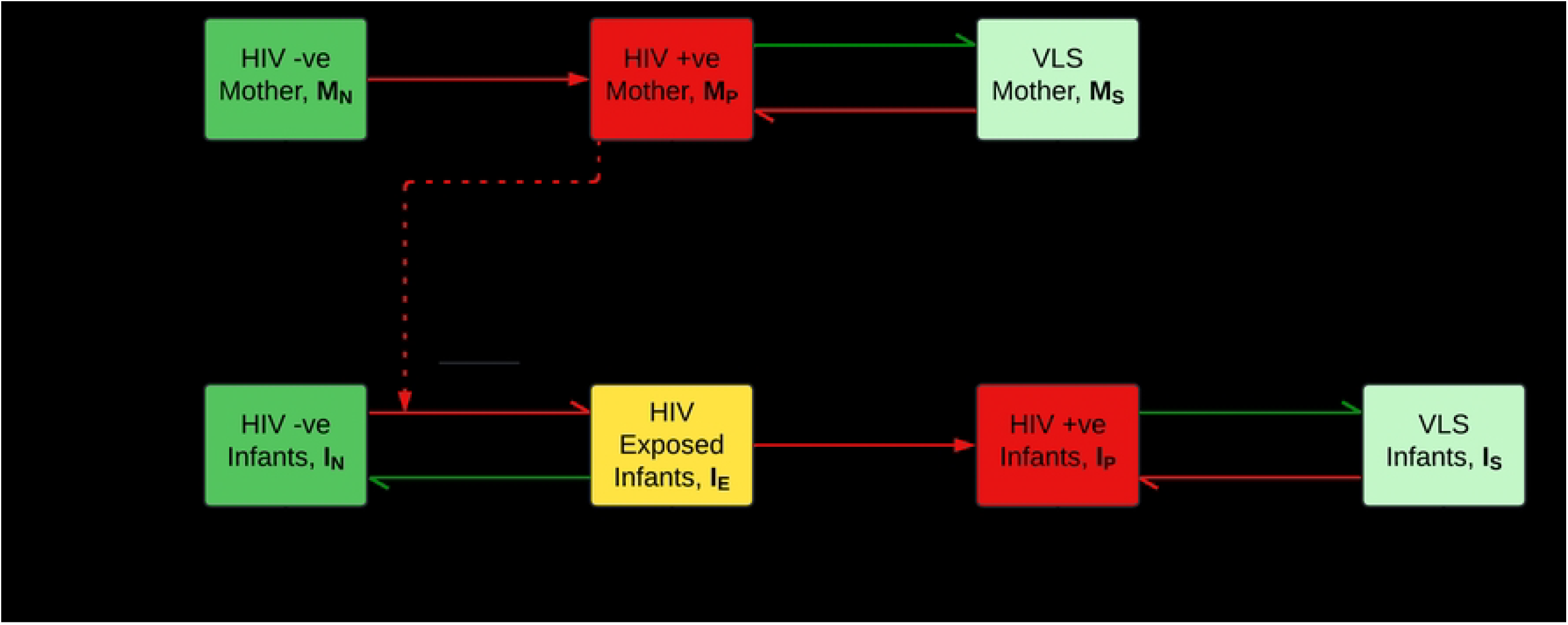

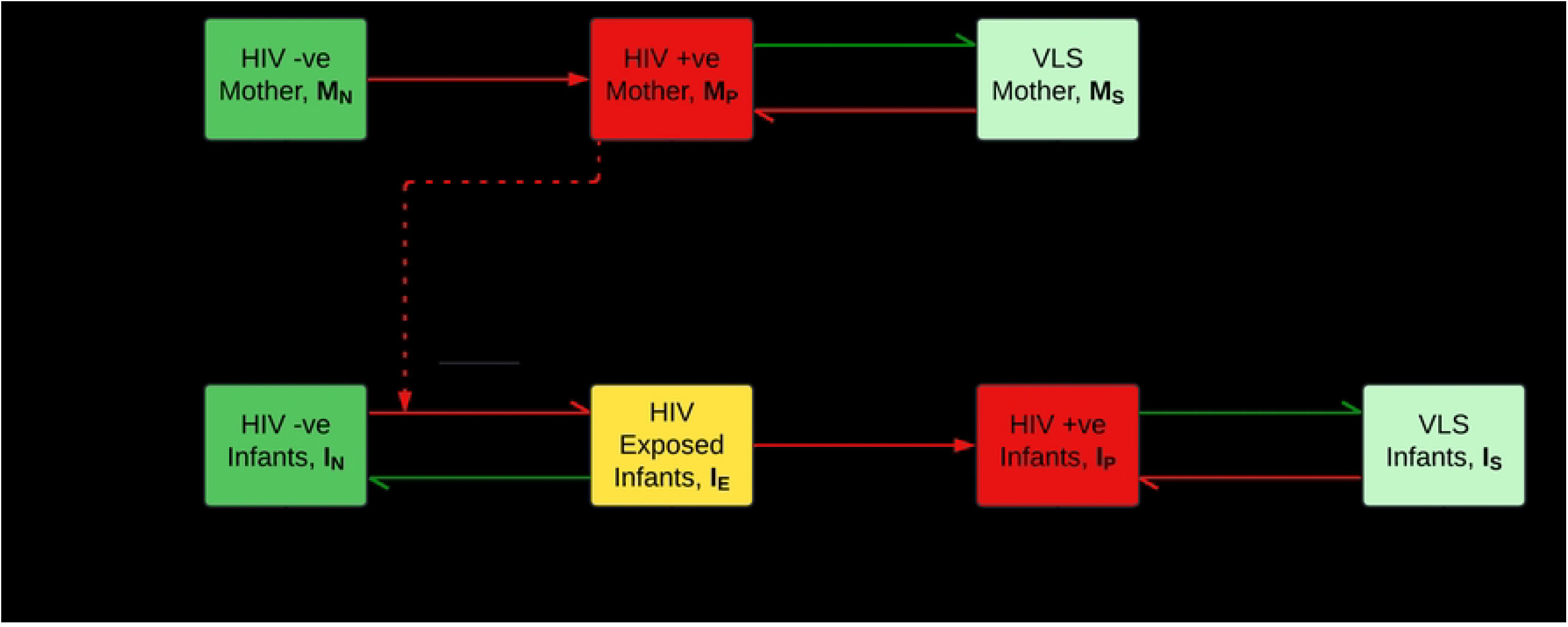

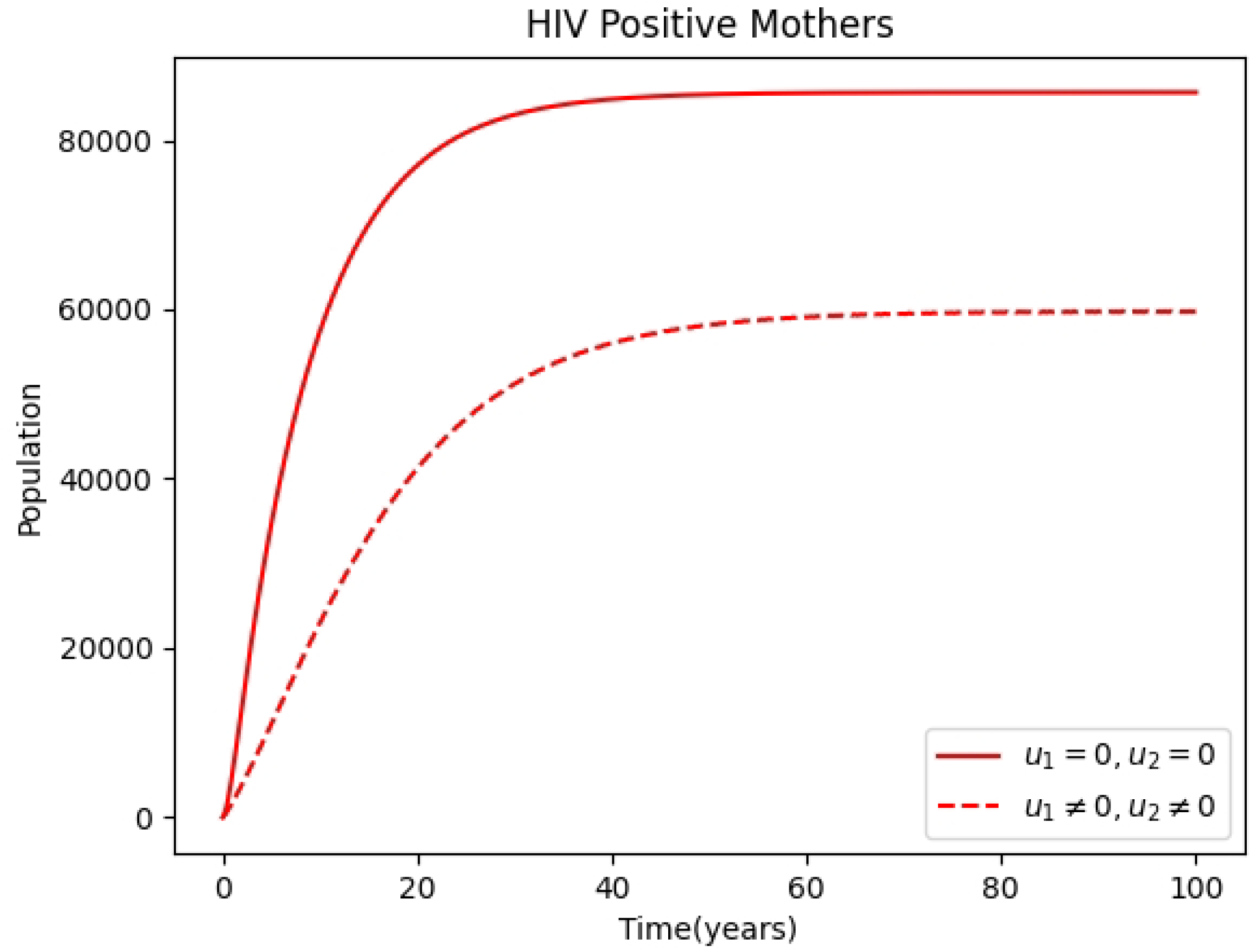

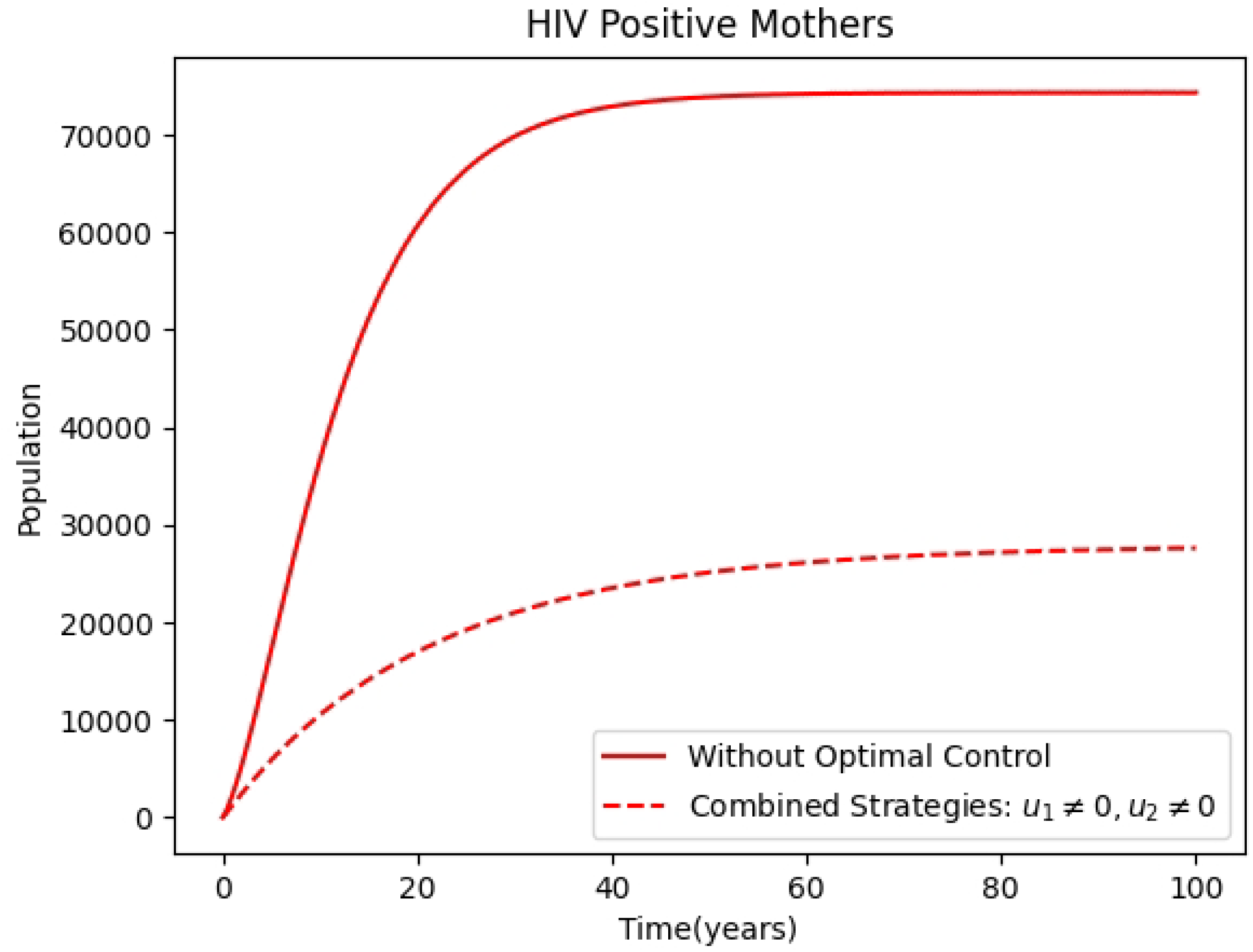

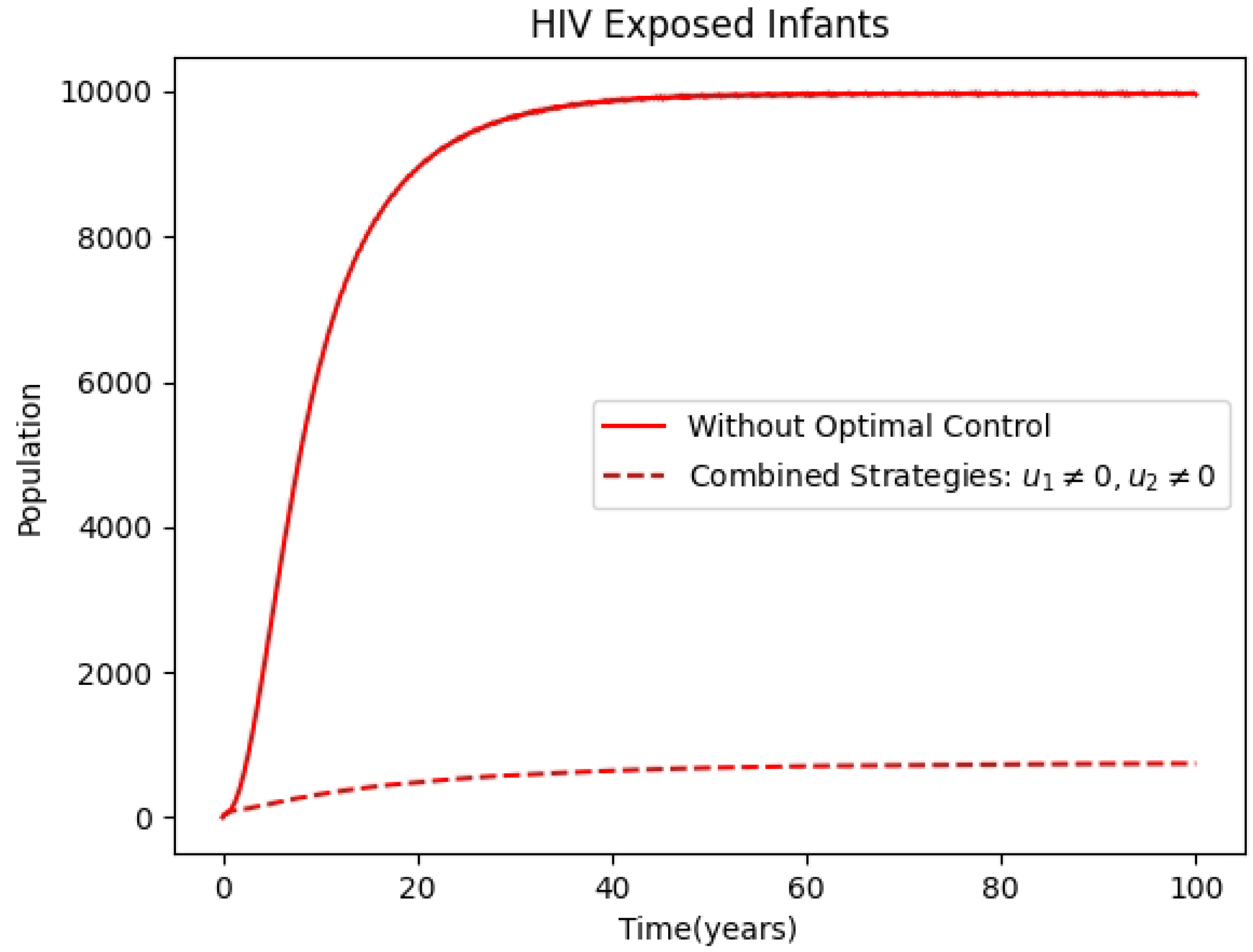

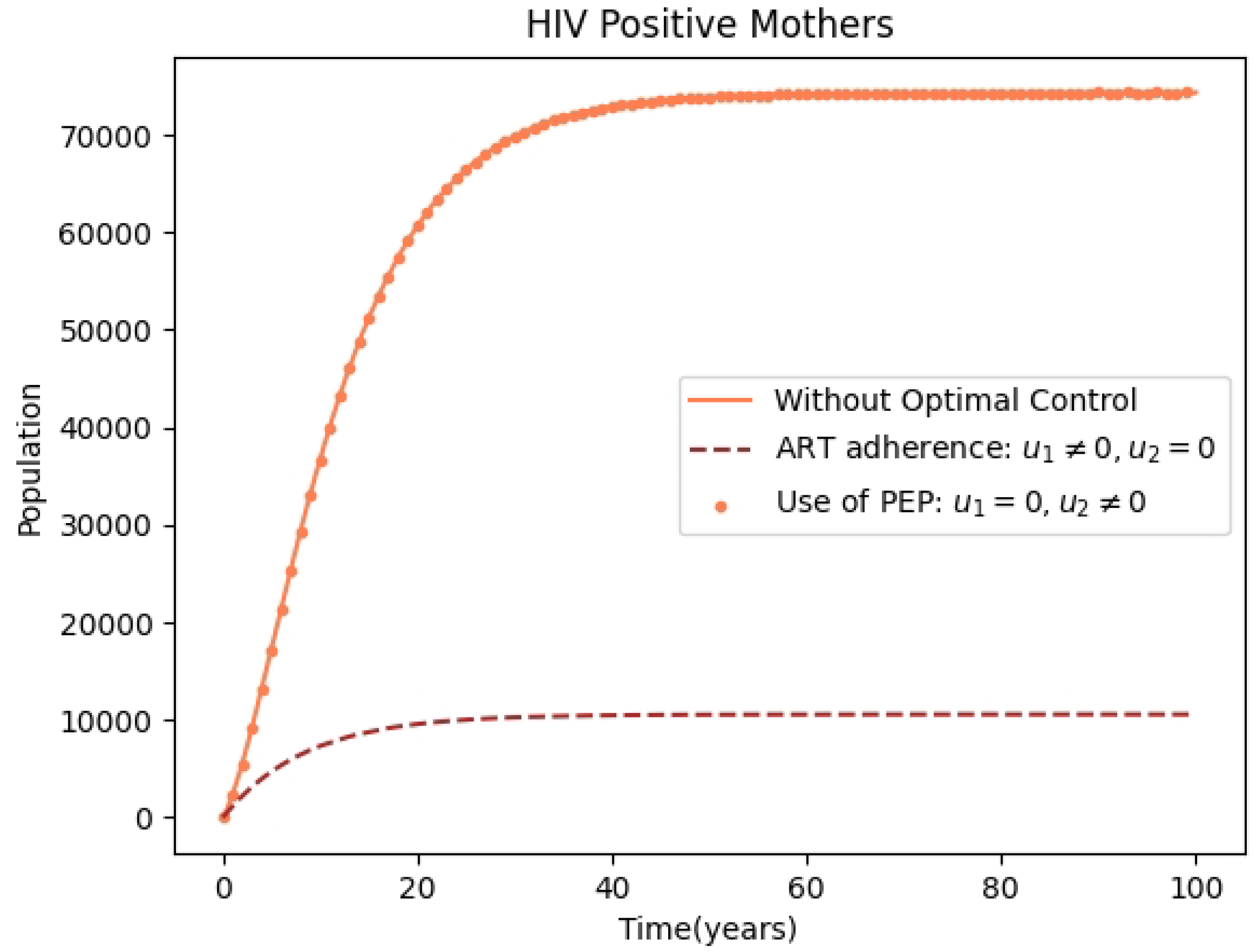

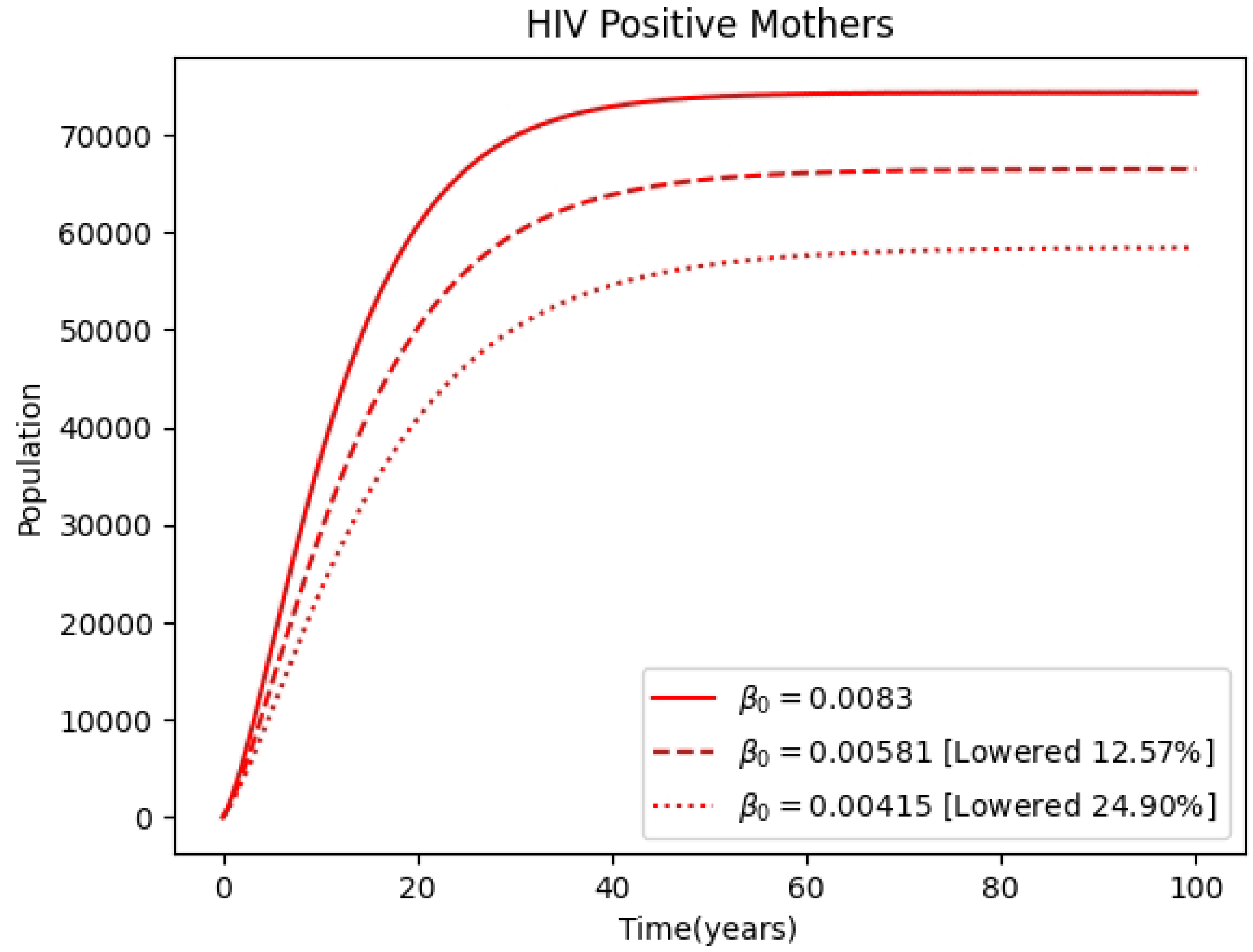

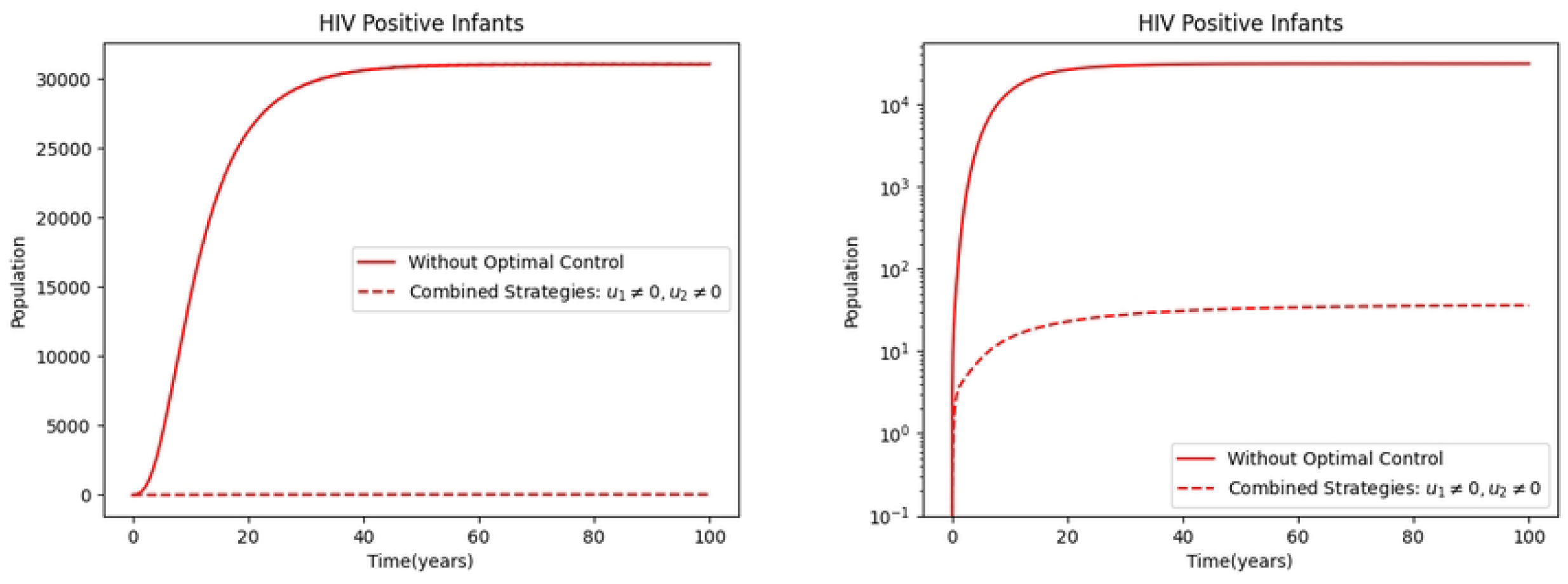

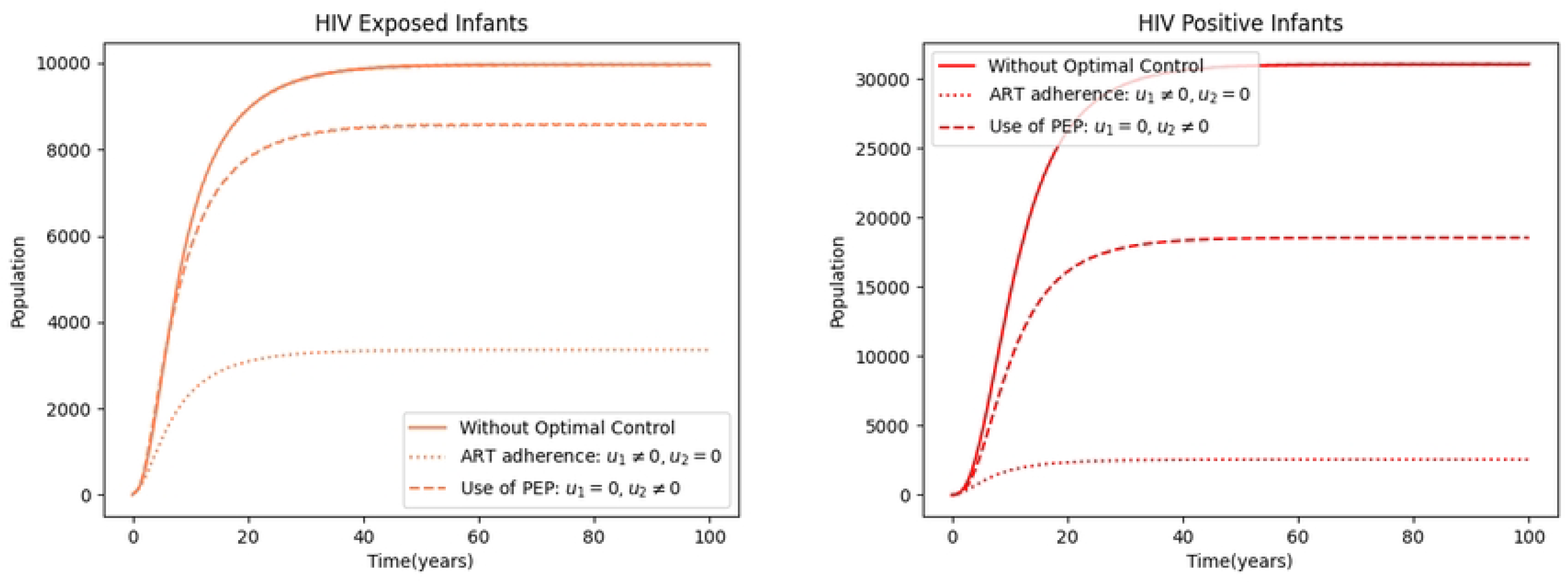

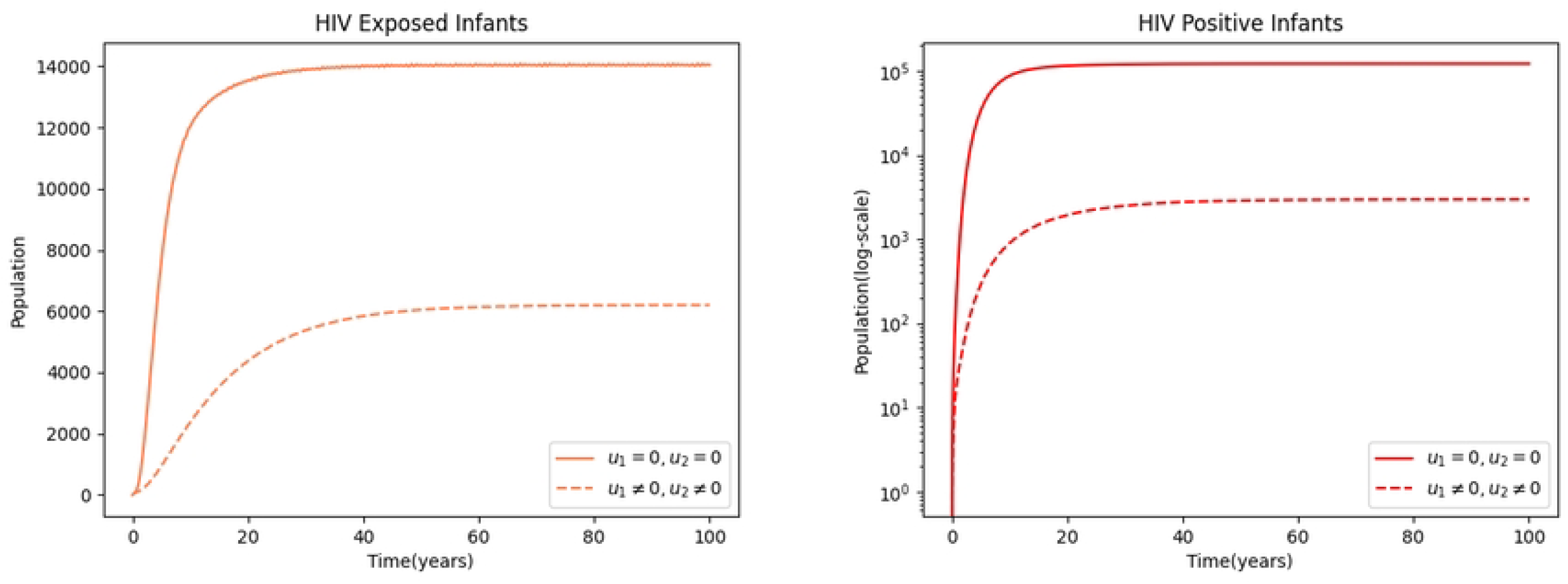

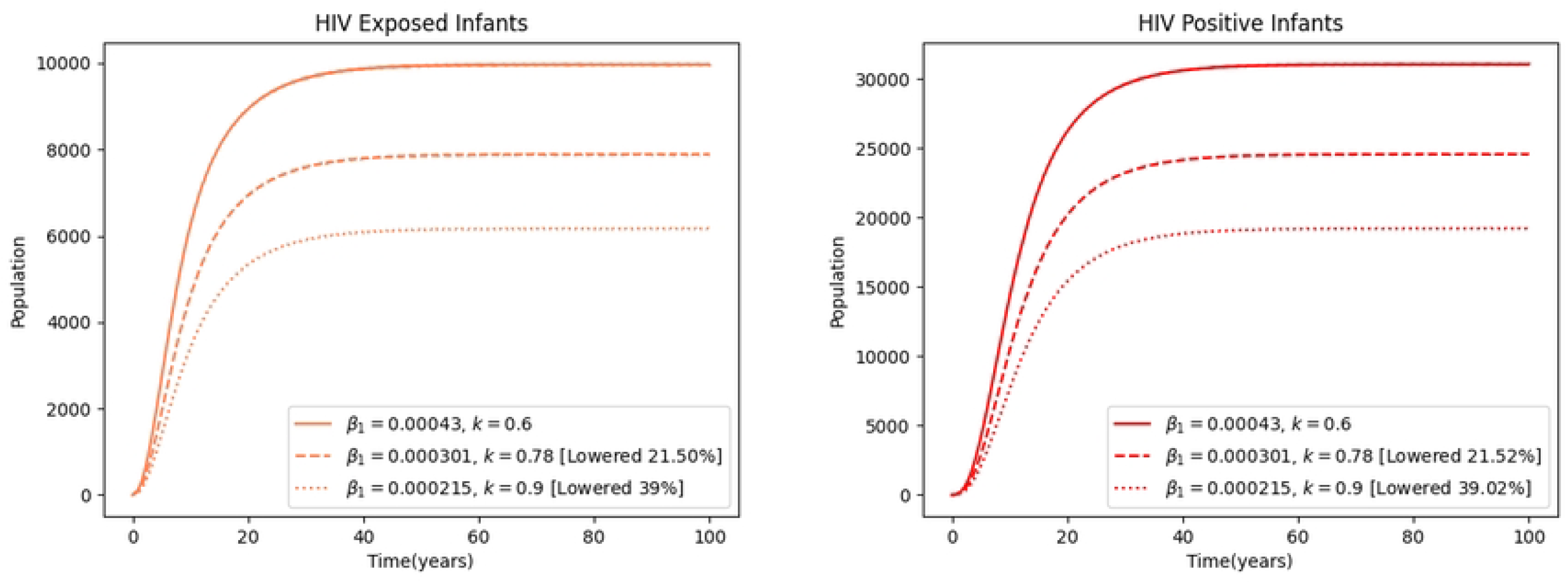

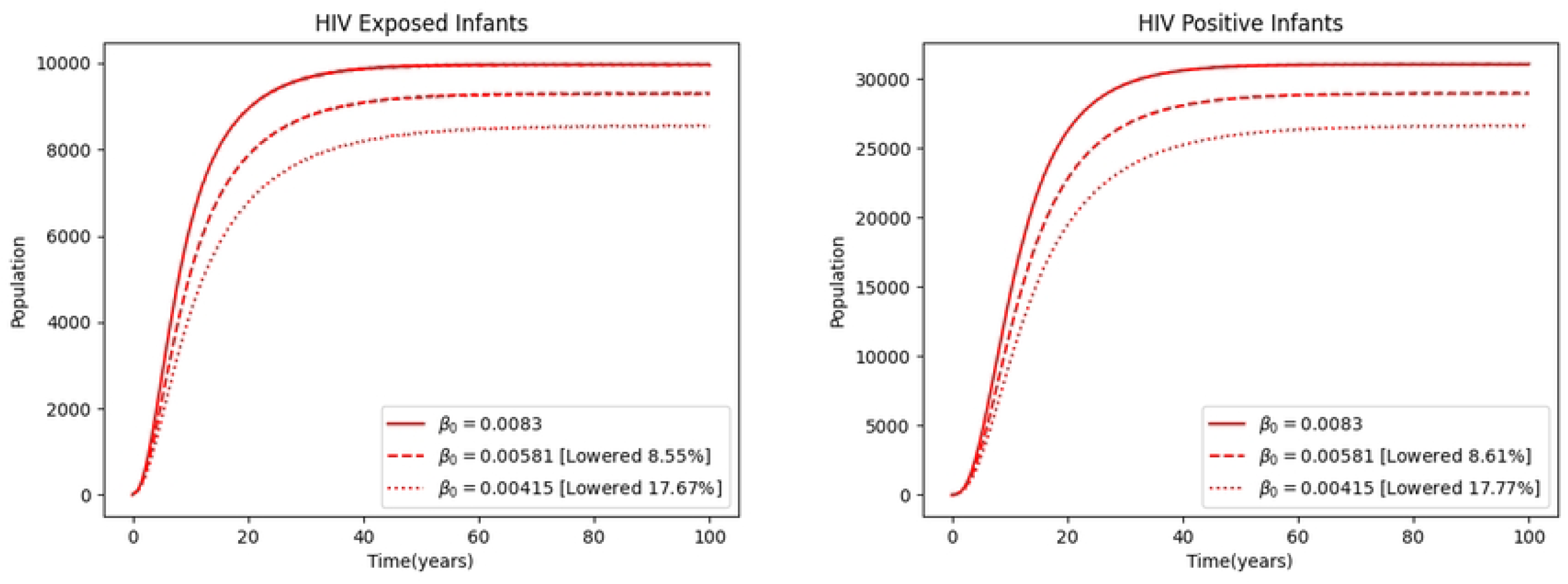

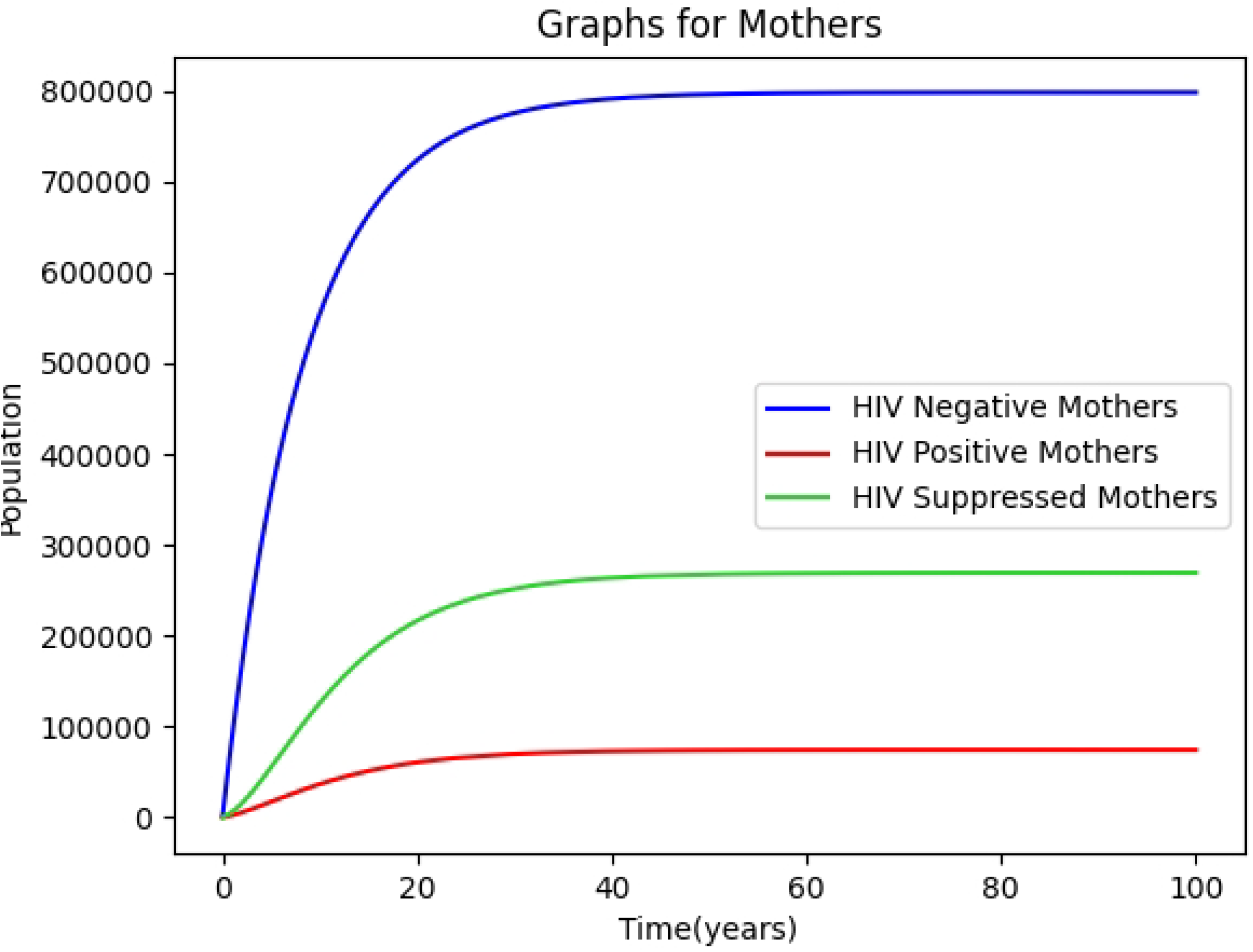

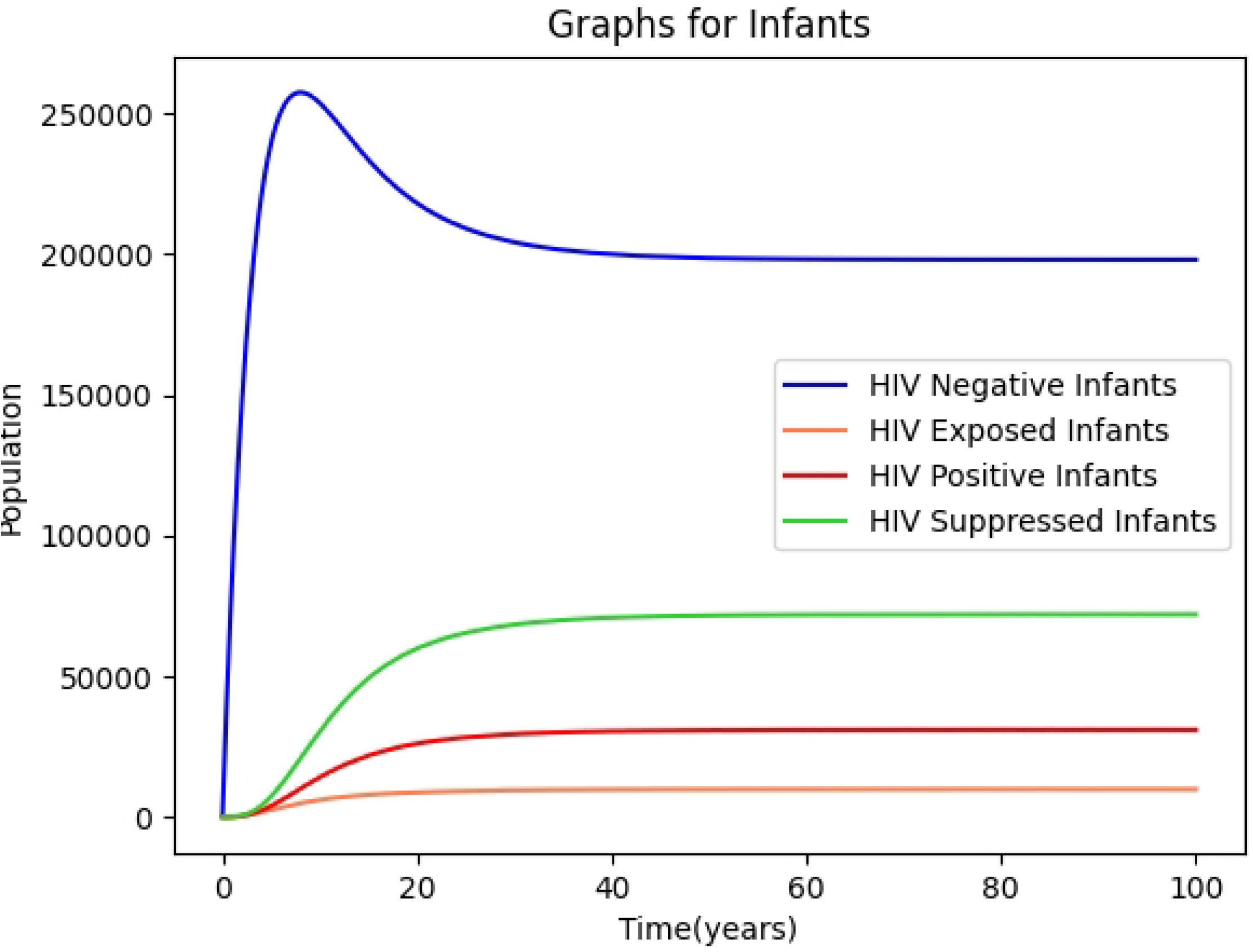

